# Large language models for generating medical examinations: systematic review

**DOI:** 10.1101/2024.01.06.24300920

**Authors:** Yaara Artsi, Vera Sorin, Eli Konen, Benjamin S. Glicksberg, Girish Nadkarni, Eyal Klang

**Affiliations:** Azrieli Faculty of Medicine, Bar-Ilan University, Zefat, Israel; Department of Diagnostic Imaging, Chaim Sheba Medical Center, Israel; Tel-Aviv University School of Medicine, Israel; DeepVision Lab, Chaim Sheba Medical Center, Israel; Division of Data-Driven and Digital Medicine (D3M), Icahn School of Medicine at Mount Sinai, New York, New York, USA; The Charles Bronfman Institute of Personalized Medicine, Icahn School of Medicine at Mount Sinai, New York, New York, USA

**Keywords:** Large language models, GPT, Multiple choice questions, Medical education, Artificial intelligence, Medical examination

## Abstract

**Purpose:** Writing multiple choice questions (MCQs) for the purpose of medical exams is challenging. It requires extensive medical knowledge, time and effort from medical educators. This systematic review focuses on the application of large language models (LLMs) in generating medical MCQs.

**Methods:** The authors searched for studies published up to November 2023. Search terms focused on LLMs generated MCQs for medical examinations. MEDLINE was used as a search database.

**Results:** Overall, eight studies published between April 2023 and October 2023 were included. Six studies used Chat-GPT 3.5, while two employed GPT 4. Five studies showed that LLMs can produce competent questions valid for medical exams. Three studies used LLMs to write medical questions but did not evaluate the validity of the questions. One study conducted a comparative analysis of different models. One other study compared LLM-generated questions with those written by humans.

All studies presented faulty questions that were deemed inappropriate for medical exams. Some questions required additional modifications in order to qualify.

**Conclusions:** LLMs can be used to write MCQs for medical examinations. However, their limitations cannot be ignored. Further study in this field is essential and more conclusive evidence is needed. Until then, LLMs may serve as a supplementary tool for writing medical examinations.

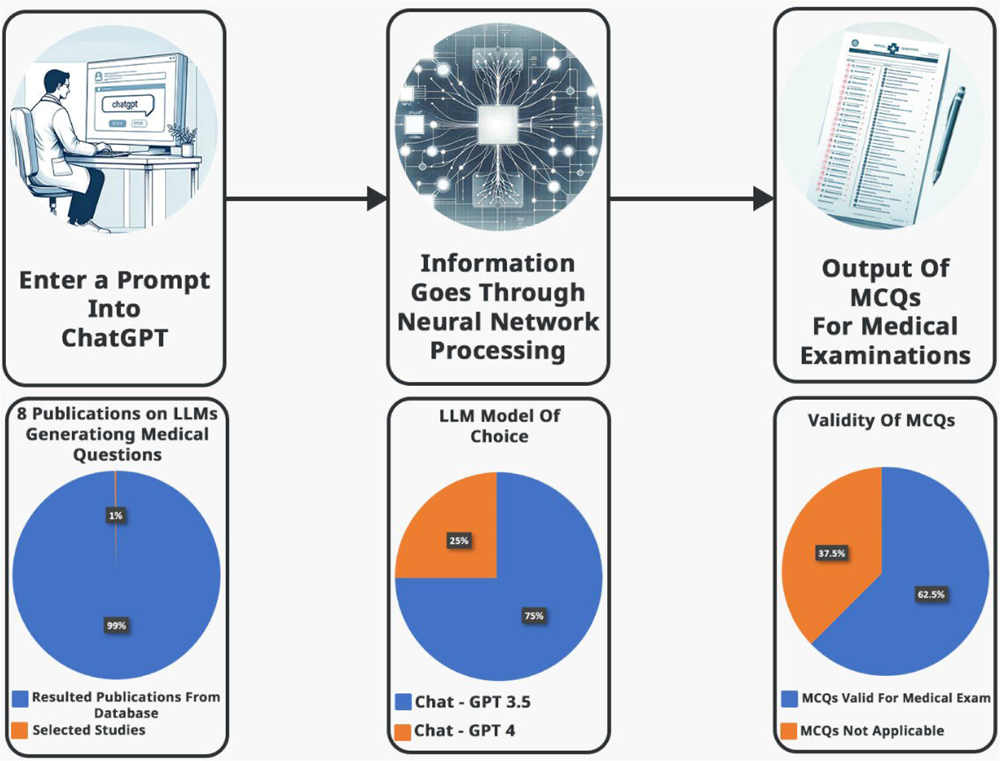

## Introduction

There is a global shortage of clinical practitioners and increasing demand for medical professionals. This need presents significant challenges in the healthcare system [1, 2, 3]. In response, the number of medical schools and students has been rising worldwide [4, 5], leading to an increase in the demand for written tests.

Creating multiple choice questions (MCQs) requires medical knowledge, conceptual integration, and avoiding potential pitfalls. For example, repeat items in examinations from year to year, or inherent imperfections called item-writing flaws (IWFs). Although at first sight IWFs may appear trivial, they can affect the way students understand and answer questions [6, 7, 8, 9]. Producing MCQs is also time consuming. Any application capable of automating this process could be highly valuable [10, 11].

Amidst these challenges, advancements in natural language processing (NLP) are constantly discussed and evaluated [12]. In particular, the introduction of OpenAI’s state-of-the-art large language models (LLMs) such as GPT-3.5 and GPT-4 [13, 14]. These models offer potential solutions to healthcare education, due to their human-like text understanding and generation, which includes clinical knowledge [15]. This could be pivotal in automating the creation of medically precise MCQs. However, automating MCQs creation introduces potential risks, as the accuracy and quality of AI generated content is still in question [16, 17].

We aimed to review the literature on LLMs’ ability to generate medical questions. We evaluated their clinical accuracy and suitability for medical examinations.

## Methods

### Literature search

On November 2nd 2023 we conducted a search identifying studies describing LLMs’ applications in generating medical questions. We searched PubMed/MEDLINE for papers with the following keywords, using Boolean operators AND/ OR: large language models; GPT; ChatGPT; medical questions; medical education; USMLE; MCCQE1; board exam; medical exam.

We also checked the references list of selected publications for more relevant papers. Sections as ‘Similar Articles’ below articles (e.g., PubMed) were also inspected for possible additional articles.

Ethical approval was not required, this is a systematic review of previously published research, and does not include any individual participant information.

Our study followed the Preferred Reporting Items for Systematic Reviews and Meta-Analyses (PRISMA) guidelines.

The study is registered with PROSPERO (CRD42023481851)

### Inclusion and exclusion process

Publications resulting from the search were initially assessed by one author (YA) for relevant titles and abstracts. Next, full-text papers underwent an independent evaluation by two authors (EK and VS).

We included full length studies describing LLMs generating medical questions. We excluded papers dating before 2023, non-English papers and non-original studies. Any study in question was discussed among all authors until reaching a unanimous agreement. (**Figure 1**.)

**Figure 1:** Flow Diagram of the Inclusion Process. Flow diagram of the search and inclusion process based on the Preferred Reporting Items for Systematic Reviews and Meta-Analyses (PRISMA) guidelines, November 2023.

Risk of bias and applicability were evaluated using the tailored QUADAS-2 tool. (**Figure 2**.)

**Figure 2:** QUADAS-2 table for potential bias and applicability. Risk of bias and applicability were evaluated using the tailored QUADAS-2 tool, November 2023.

## Results

### Study selection and characteristics

The initial literature search resulted in 838 articles. Eight studies met our inclusion criteria (**Figure 1**). Most studies were retrospective: 6/8 (75%). One study is cross-sectional and one study is prospective. Most studies used Chat-GPT (3.5 or 4) as an AI model of choice, other models evaluated included Microsoft’s Bing and Google’s Bard. The MCQs were produced with varying parameters (**Table 1**). Overall 5/8 (62.5%) studies demonstrated valid MCQs. 6/8 (75%) of the studies utilized the latest version Chat-GPT 4 (**Figure 3**.)

**Figure 3:** Illustration of multiple-choice questions (MCQs) generation and summary of preliminary results. The images in the upper row illustrate the MCQs generation process via a large language model. The images in the bottom row showcase preliminary data results, November 2023.

**Table 1:** Summary of the articles in the literature that applied AI for generating medical questions, November 2023.

### Descriptive summary of results

Cheung et al. [18] were the first, and so far, the only study to compare LLM to humans in MCQs writing. Chat-GPT 3.5 plus generated the MCQs. The reference for the prompt were two standard undergraduate medical textbooks: Harrison’s Principles of Internal Medicine the 21th edition for medicine [19], and Bailey and Love’s Short Practice of Surgery 27th Edition for surgery [20]. Only four choices were given per question. Also, only text and knowledge-based questions were generated. No modification to the MCQs was allowed after generation. Chat-GPT 3.5 performed relatively well in the task. The overall time required for the AI to generate 50 MCQs was 21 minutes. This is about 10% of the total time human writing required (211 min). However, the questions written by humans were far better. Both in terms of quality and validity, outperforming the AI in a total score of 30 (60%) eligible MCQs (**Table 2**).

**Table 2:** Summary of key parameters investigated in each study, November 2023.

Klang et al. [21] performed blind assessment of the generated questions. They did not disclose to the evaluators whether the MCQs origin was AI. At first, they asked Chat-GPT 4 to create MCQs on the topic of internal medicine. They used as reference (few-shot learning) a former exam of the same subject. The MCQs had four possible answers, with the correct answer marked with an asterisk. At first, the generated MCQs were short with no clinical background. This required a second prompting of the AI model, specifically requesting the AI to create MCQs with clinical history. The study showed promising results, with the majority of MCQs deemed valid as exam questions (**Table 2**).

In a cross-sectional study, Agarwal et.al [22] compared different LLMs. They compared Chat-GPT 3.5/Bard/Bing in MCQs generating capability. They used as reference the 11-module curriculum for physiology, created by The Indian National Medical Commission (NMC). The authors requested in the prompt to Generate five difficult reasoning-based MCQs, fitting levels of Bachelor of Medicine, and Bachelor of Surgery (MBBS). Chat-GPT’s generated MCQs were significantly more valid than the other AI tools examined in the study. However, the difficulty level was lower compared to Bard and Bing (**Table 2**).

Ayub et al. [23] focused on medical board examination for Dermatology. They utilized Chat-PDF to upload entire PDF files into a ChatGPT 3.5 portal. The reference used was “Continuing medical education” (CME) articles, taken from the *Journal of the American Academy of Dermatology* (JAAD). This reference is considered high-yield review material for the American Board of Dermatology Applied Exam (ABD-AE). This study’s prompt was not detailed in the paper. The three parameters to evaluate the MCQs were accuracy, complexity, and clarity. Only 16 (40%) of the generated questions were applicable (**Table 2**). The rest were unclear 9 (22%), inaccurate 5 (13%) or had low complexity 10 (25%) (**Table 3**).

Sevgi et al. [24] asked Chat-GPT 3.5 to prepare three questions with answers and explanations at a level appropriate for a neurosurgery board exam. There was no independent evaluation of the MCQs.

Han et al. [25] instructed Chat-GPT 3.5 to write three MCQs, each containing clinical background and lab values. Each time they requested Chat-GPT to rephrase the question. First, for a different correct answer and then for an increased level of difficulty. There was no independent evaluation of the MCQs.

Totlis et al. [26] asked Chat-GPT 4 to generate MCQs on the topic of anatomy. In the prompt they requested increasing difficulty and matching correct pairs. There was no independent evaluation of the MCQs.

Biswas [27] requested in the prompt to prepare MCQs for USMLE step 1 exam. There was no independent evaluation of the MCQs.

All studies presented some faulty questions that were deemed inappropriate for medical exams. Some questions required additional modifications in order to qualify (**Table 3**). We included examples from each study, demonstrating valid MCQs as well as faulty MCQs for various reasons (**Table 4**.)

**Table 3:** Summary of faulty questions generated by the AI, November 2023.

**Table 4:** Examples from studies of multiple-choice questions generated by AI. Both valid and faulty, November 2023.

## Discussion

In this study we explored LLMs’ applicability in generating medical questions. Specifically, multiple choice questions (MCQs) for medical examinations.

MCQs are an essential component of medical exams, used in almost every aspect of medical education [8, 9]. Yet, they are time consuming and expensive to create [28]. The possibility of AI generated questions can provide an important opportunity for the medical community and transform the way written tests are generated. Using LLMs to support these tasks can potentially save time, money and reduce burnout. Especially in a system already sustaining itself on limited resources [29].

### AI benefits

Physician burn-out, poor mental health and growing personal distress are constantly studied [30]. However, academic physicians experience a unique set of additional challenges, such as increased administrative work, less time with patients and increased clinical responsibilities. As a result, they have less time for traditional academic pursuits such as research and education [31, 32, 33]. In the famous words of Albert Einstein: “Bureaucracy is the death of any achievement”.

AI can potentially relieve medical educators from tiresome bureaucracy and administrative work, allowing them to focus on the areas that they view as most personally meaningful and avoid career dissatisfaction [32, 34].

According to Bond et al. another possible application of AI in medical education is grading patients notes. This can provide additional formative feed-back for students in the face of limited faculty availability [35]. Moreover, AI can assist medical students by creating personalized learning experience, while accessing current up-to-date information [36]. These are only a few examples, as every day new tasks improved by AI are discovered.

### AI drawbacks

Nowadays, AI continues to evolve, becoming more integrated in various medical fields [37]. The probability of revolutionizing the healthcare world seems inevitable. AI performance is fast, efficient and with what seems like endless data resources [38]. In almost every study we reviewed, LLMs’ execution was more than satisfactory with the consensus that AI is capable of producing valid questions for medical exams. However, while these models show promise as an educational tool, their limitations must be acknowledged.

One notable limitation is a phenomenon known as “Hallucination” [39]. This occurs when generative AI misinterprets the given prompt, resulting in outputs that lack logical consistency. When relying on AI for quality MCQs writing, this phenomenon is concerning. Furthermore, AI ability to integrate contextual and sensory information is still not fully developed. Currently, AI cannot understand non-verbal cues or body language. Also, bias in data and inaccuracy is troubling [40, 41].

Another consideration is the logistics necessary to implement AI in healthcare and education. New technologies require training, commitment and investment in order to be maintained and managed in a sustainable way. Such a process can take time and energy [42]. Moreover, implementing new technology increases concern for privacy and data security [43, 44].

Patients’ data is sensitive and often a target for cyber-attacks [45].

Equally important limitation of AI integration in healthcare is accountability. AI “black box” refers to the “knowledge within the machine”. The internal workings of the system are invisible to the user. Healthcare staff use the AI, write the input and receive the output. But, the system’s code or logic cannot be questioned or explained [46].

Additional aspect to consider is the longstanding concern of AI replacing human jobs [47]. This could cause a dislike and resistance to AI integration. This notion is unlikely in the near future. But, distrust in AI technology is yet another challenge to its implementation [48].

But maybe the biggest concern of AI application in medical education is impairing students’ critical thinking. According to Van de Ridder et al., self-reflection and criticism are crucial for a medical student’s learning process and professional growth. In a reality where a student can delegate to Chat-GPT tasks such as writing personal reflection or learning experiences, the students deny themselves of the opportunity to self-reflect and grow as physicians [49].

It is imperative to take into consideration those significant shortcomings and challenges. AI should be used wisely and responsibly while integrating it into medical education.

### MCQs creation

For each study we examined the process of crafting the MCQs. We noticed a wide range of approaches to writing the prompts. In some studies, additional modifications took place in order to improve the validity of the questions. This emphasizes the importance and sensitivity of prompts. Prompt-engineering may be a task that requires specific training, so that the prompt is phrased correctly and the MCQs quality is not impaired.

### Limitations

Our review has several limitations. Most of the studies are retrospective in nature. Due to heterogeneity in study design and data, a meta-analysis was not performed. None of the questions were image or graph based, which is an integral part of medical exams. Three studies did not base their prompt on a valid medical reference, such as previous exams or approved syllabus. Also, the three studies did not evaluate the questions after they were generated. Two studies were at high risk of bias.

Additional studies will be needed to further solidify the usefulness of AI tools, especially in generating competent medical exam questions.

Lastly, we limited our search to PubMed/MEDLINE. We did so due to its relevance in biomedical research. We recognize this choice narrows our review’s scope. This might exclude studies from other databases, possibly limiting diverse insights.

## Conclusion

AI-generated MCQs for medical exams are feasible. The process is fast and efficient, demonstrating great promise in the future of medical education and exam preparation. However, their use warrants cautious and critical evaluation. Awareness of AI limitations is imperative in order to avoid misuse and deterioration of medical education quality. Currently, further research in this field is needed. Until more advancements are achieved, AI should be viewed as a powerful tool best utilized by experienced professionals.

## Disclosure statement

### Disclosure of interest

The authors report there are no competing interests to declare

## Additional information

### Funding

The author(s) reported there is no funding associated with the work featured in this article.

## Data Availability

All data produced in the present work are contained in the manuscript

## Acknowledgments

None

## Disclaimer

None

## Notes

### Competing Interest Statement

The authors have declared no competing interest.

### Funding Statement

This study did not receive any funding

