## Supplementary material for "Large language models for generating medical examinations: systematic review": Tables 1-3

**Table 1:** A summary of the articles in the literature that applied AI for generating medical questions, November 2023

| **Study** | **Author** | **Month** | **Journal** | **Study design** | **AI tool** |
| --- | --- | --- | --- | --- | --- |
| 1 | Sevgi et.al | April | Springer Link | Retrospective | Chat-GPT 3.5 |
| 2 | Biswas | May | Springer Link | Retrospective | Chat-GPT 3.5 |
| 3 | Agarwal et.al | June | Cureus | Cross-sectional study | Chat-GPT,  Bard, Bing |
| 4 | Ayub et.al | August | Cureus | Retrospective | Chat-GPT 3.5 |
| 5 | Cheung et.al | August | PLOS ONE | Prospective | Chat-GPT 3.5 plus |
| 6 | Totlis et.al | August | Springer Link | Retrospective | Chat-GPT 4 |
| 7 | Han et.al | October | Taylor & Francis | Retrospective | Chat-GPT 3.5 |
| 8 | Klang et.al | October | Springer Nature | Retrospective | Chat-GPT 4 |

**Table 2:** A summary of key parameters investigated in each study, November 2023

| **Author** | **No. of MCQs** | **Tested vs Human** | **Medical Field** | **Questions Evaluated By** | **Performance**  **Scores** |
| --- | --- | --- | --- | --- | --- |
| Sevgi et.al | 3 | No | Neurosurgery | Evaluated by the author according to current literature | 2 (66.6%) of the questions were accurate |
| Biswas | 5 | No | General | N/A | N/A |
| Agarwal et.al | 320 | No | Medical Physiology | 2 Physiologists | **p value validity of < 0.001 for:**  Chat-GPT vs. Bing < 0.001  Bard vs. Bing < 0.001 **p value of difficulty 0.006**  ChatGPT vs Bing 0.010  ChatGPT vs Bard 0.003 |
| Ayub et.al | 40 | No | Dermatology | 2 board certified dermatologists | 16 (40%) of questions valid for exams |
| Cheung et.al | 50 | Yes | Internal Medicine/Surgery | 5 International medical experts and educators | **Overall performance:**  AI score 20 (40%) vs. Human score 30 (60%)  Mean difference -0.80 ± 4.82 **Total time required:**  AI 20 min 25 sec vs. Human 211 min 33 sec |
| Totlis et.al | 18 | No | Anatomy | N/A | N/A |
| Han et.al | 3 | No | Biochemistry | N/A | N/A |
| Klang et.al | 210 | No | Internal Medicine Surgery Obstetrics & Gynecology Psychiatry Pedicatrics | 5 Specialist physicians in the tested fields | **Problamatic questions by field:**  Surgery 30%  Gynecology 20%  Pediatrics 10%  Internal medicine 10%  Psychiatry 0% |

**Table 3:** A summary of faulty questions generated by the AI, November 2023

| **Author** | **Medically Irrelevant Questions** | **Invalid for Medical  Exam** | **Inaccurate/Wrong Question** | **Inaccurate/Wrong Answer or Alternative answers** | **Low**  **Difficulty**  **Level** |
| --- | --- | --- | --- | --- | --- |
| Sevgi et.al | N/A | N/A | N/A | 1 (33.3%) | N/A |
| Biswas | N/A | N/A | N/A | N/A | N/A |
| Agarwal et.al | N/A | Highly valid | N/A | V/A | Somewhat difficult |
| Ayub et.al | 9 (23%) | 24 (60%) | 5 (13%) | 5 (13%) | 10 (25%) |
| Cheung et.al | 32 (64%) | 28 (56%) | 32 (64%) | 29 (58%) | N/A |
| Totlis et.al | N/A | 8 (44.4%) | N/A | N/A | 8 (44.4%) |
| Han et.al | N/A | N/A | N/A | N/A | 3 (100%) |
| Klang et.al | 2 (0.95%) | 1 (0.5%) | 12 (5.7%) | 14 (6.6%) | 2 (0.95%) |

**Table 4:** Examples from studies showcasing valid and faulty MCQs, November 2023

| **Study** | **Correct MCQs** | **Faulty MCQs** | | | |
| --- | --- | --- | --- | --- | --- |
|  |  | **More than one correct answer** | **Inaccurate MCQs** | **Low complexity** | **Unclear** |
| **Sevgi et al.** | A 45- year old woman presents to the clinic with a 6-month history of gradually worsening left-sided weakness and numbness. She reports that she has also noticed some changes in her vision, including difficulty reading small print. Physical examination reveals left hemiparesis and left homonymous hemianopsia. An MRI of the brain reveals a large, enhancing mass in the left parietal lobe. What is the most appropriate management for this patient?   A) Observation and follow-up imaging  B) Stereotactic radiosurgery  C) Chemotherapy  D) Craniotomy and resection of the mass  E) Whole brain radiation therapy | Which of the following vessels is NOT a component of the Circle of Willis?   A) Anterior cerebral artery.  B) Posterior communicating artery.  C) Middle cerebral artery.  D) Vertebral artery.  E) Superior cerebellar artery.   **→ Correct answer:  D and E** | N/A | N/A | N/A |
| **Biswas et al.** | Which of the following medications is contraindicated in patients with a history of sulfonamide allergy?   A) Furosemide  B) Hydrochlorothiazide   C) Spironolactone  D) Triamterene  E) Amiloride | N/A | N/A | N/A | N/A |
| **Agarwal et al.** | Which of the following processes requires the expenditure of energy to transport molecules across the cell membrane?   A) Passive transport  B) Osmosis  C) Simple diffusion  D) Active transport | N/A | N/A | N/A | N/A |
| **Ayub et al.** | A 40-year-old female presents with a new onset of pruritic, scaly plaques on her scalp and forehead. She has a history of rheumatoid arthritis and has been on methotrexate therapy for the past year. What is the most appropriate next step in management?   A) Increase the dose of methotrexate  B) Switch to cyclosporine therapy  C) Discontinue methotrexate and initiate topical corticosteroids.  D) Perform a skin biopsy | N/A | N/A | Which of the following is a characteristic feature of melanoma?   A) Uniform color  B) Smooth bordes  C) Symmetry  D) Irregular pigmentation | Which technique correctly diagnosed lesions 82% in differentiating biopsy-proven DN from thin melnomas?   A)Qualitative pattern analysis  B) ABCD rule of dermoscopy  C) The 7-point checklist  D) All of the above |
| **Cheung et al.** | N/A | N/A | | | |
| **Totlis et al.** | What is the anatomical term for the socket in the pelvic bone where the femur articulates?   A) Acetabulum  B) glenoid cavity  C) foramen magnum  D) fossa ovalis | N/A | N/A | N/A | N/A |
| **Han et al.** | A 35-year-old woman presents with a history of obesity and type 2 diabetes. She is found to have a triglyceride level of 800 mg/dL and an HDL level of 25 mg/dL. What is the most likely cause of her hyperlipidemia?   A) Familial hypertriglyceridemia  B) Familial combined hyperlipidemia  C) Metabolic syndrome  D) Secondary hyperlipidemia | N/A | N/A | N/A | N/A |
| **Klang et al.** | Which of the following is a negative symptom of schizophrenia?   A) Hallucinations  B) Delusions  C) Anhedonia  D) disorganized speech | N/A | **Example A:** 38-year-old woman with irregular menses as postmenopausal.  **Example B:** Abdominal complaint of a male was questioned, and the optional answers included ectopic pregnancy and ovarian cyst rupture. | N/A | N/A |
