## Supplementary material for "Large language models for generating medical examinations: systematic review": Figures 1-2

**Figure 1:** Flow diagram of the search and inclusion process in the study

**Identification of studies via databases and registers**

Records removed *before screening*:

Duplicate records removed (n = 0)

Records marked as ineligible by automation tools (n = 0)

Records removed for other reasons (n = 150)

Records identified from*:

Databases (n = 838)

Registers (n = 0)

**Identification**

Records screened

(n = 688)

Records excluded**

(n =613)

Reports sought for retrieval

(n =75)

Reports not retrieved

(n = 3)

**Screening**

Reports assessed for eligibility

(n =72)

Reports excluded:

Reason 1 not English (n = 1)

Reason 2 irrelevant (n =60)

Reason 3 not original study (n =3)

etc.

Studies included in review

(n = 8)

Reports of included studies

(n = 0)

**Included**

**Figure 2.** Risk of Bias and Applicability Judgments in QUADAS-2

|  | **RISK OF BIAS** | | | |  | **APPLICABILITY CONCERNS** | | |
| --- | --- | --- | --- | --- | --- | --- | --- | --- |
|  | **PATIENT SELECTION** | **INDEX TEST** | **REFERENCE STANDARD** | **FLOW AND TIMING** |  | **PATIENT SELECTION** | **INDEX TEST** | **REFERENCE STANDARD** |
| Sevgi et al. | **■** | **?** | **▲** | **■** |  | **▲** | **▲** | **■** |
| Biswas | **■** | **?** | **■** | **■** |  | **■** | **■** | **■** |
| Agarwal et al. | **■** | **▲** | **▲** | **■** |  | **■** | **■** | **■** |
| Ayub et al. | **■** | **■** | **▲** | **■** |  | **■** | **■** | **■** |
| Cheung et.al | **■** | **■** | **▲** | **■** |  | **■** | **■** | **■** |
| Totlis et al. | **▲** | **▲** | **▲** | **■** |  | **■** | **■** | **▲** |
| Han et al. | **▲** | **▲** | **▲** | **?** |  | **▲** | **■** | **■** |
| Klang et al. | **■** | **■** | **■** | **▲** |  | **■** | **■** | **■** |

| Low Risk | **■** |
| --- | --- |
| High Risk | **▲** |
| Unclear Risk | **?** |
